# Effect, equity and costs of an integrated and decentralised intervention to improve access to primary care for skin diseases: a prospective before-and-after study in south-west Ghana

**DOI:** 10.64898/2026.03.23.26348798

**Authors:** Stefan Witek-McManus, Richard Adjei Akuffo, Jacob Novignon, Daniel Okyere, Ruth Dede Tuwor, Edmond Kwaku Ocloo, Emmanuel Kyei Afreh, Emmanuel Boakye Boateng Okyere, Abigail Agbanyo, Abubakar Amadu, Jojo Cobbinah, Abdul Samad Akate, Adelaide Fokuoh-Boadu, Miriam Gborglah, Adwoa Asante-Poku, Eric Koka, Collins Stephen Ahorlu, Tara Mtuy, Jennifer Palmer, Yaw Ampem Amoako, Michael Marks, Catherine Pitt, Stephen L. Walker, Dorothy Yeboah-Manu, Richard O. Phillips, Rachel L. Pullan

**Affiliations:** Department of Disease Control, Faculty of Infectious and Tropical Diseases, London School of Hygiene and Tropical Medicine, London, UK; Noguchi Memorial Institute for Medical Research, University of Ghana, Accra, Ghana; Centre for Social Policy Studies, University of Ghana, Accra, Ghana; Kumasi Centre for Collaborative Research in Tropical Medicine, Kwame Nkrumah University of Science and Technology, Kumasi, Ghana; Department of Sociology and Anthropology, University of Cape Coast, Cape Coast, Ghana; Atwima Mponua District Health Directorate, Nyinahin, Ghana; Department of Clinical Research, Faculty of Infectious and Tropical Diseases, London School of Hygiene and Tropical Medicine, London, UK; Department of Global Health and Development, Faculty of Public Health and Policy, London School of Hygiene and Tropical Medicine, London, UK; Hospital for Tropical Diseases, University College Hospital, London, UK; School of Medical Sciences, Kwame Nkrumah University of Science and Technology, Kumasi, Ghana

## Abstract

**Background:** Global guidelines recommend strengthening and integrating health services for skin diseases, yet evidence for strategies remains scarce. We evaluated a decentralised approach to the care of skin disease through integration within routine primary care, by assessing uptake, equity, alignment with skin disease burden and associated treatment costs.

**Methods and findings:** A before-and-after intervention study was conducted across all 17 public health facilities in Atwima Mponua district, Ghana from November 2023 to September 2024 (intervention period). We analysed routine health facility records to compare uptake of primary care for skin diseases pre-intervention (January to October 2023) and during the intervention. We assessed the burden of skin disease through a community-based two-stage cross-sectional dermatological survey, and estimated patient and provider costs for skin disease through post-care questionnaires and health facility surveys. We compared uptake to disease burden and assessed catastrophic expenditure and factors associated with higher treatment costs. Uptake of primary care for skin disease doubled during the intervention period relative to the pre-intervention period (adjusted incidence rate ratio (aIRR) 2.0, 95% CI 1.92–2.09), with greatest increases amongst school-age children (aIRR 2.70, 2.46–2.97) and individuals residing within very rural communities (aIRR 2.79, 2.47–3.15). Amongst 42,801 individuals surveyed, odds of any skin disease were greater amongst males (adjusted odds ratio (aOR) 1.25; 1.13–1.38), pre-school age children (aOR=1.80, 1.61–2.80), and residents of very rural communities (aOR=1.68, 1.09–2.61). Males and school-age children remained underrepresented amongst those who sought care during the intervention period relative to those diagnosed during the survey. Amongst patients seeking care for skin NTDs and complex wounds, 4% experienced catastrophic expenditure, driven largely by costs prior to visiting an intervention health facility.

**Conclusions:** Greater integration within primary care substantially increased uptake of care for skin disease, but populations at greatest risk remained underrepresented amongst those accessing care. These findings highlight the need for deliberate strategies to address persistent barriers to care, with lessons for integration efforts across primary health systems.

**AUTHOR SUMMARY:** *Why was this study done?:*

- Skin diseases are a common and major cause of poor health, but have historically received limited policy attention and remained a low public-health priority.
- Global recommendations have recently called for strengthening and integration of care for skin diseases into routine health services, but evidence for what is effective, fair and affordable remains very limited.
- We tested a new, comprehensive intervention in which health services for skin diseases were integrated into public primary health care in Atwima Mponua district, Ghana for one year.

*What did the researchers do and find?:*

- We compared the number of individuals who had sought care and were diagnosed with a skin disease across all public health facilities during the intervention period, as compared to the year immediately before (pre-intervention period). We also conducted detailed questionnaires with patients who received care, and a large community-based survey of skin diseases at the end of the intervention period.
- We saw that the number of skin disease cases doubled during the intervention period, with the greatest relative increases amongst school-age children (relative to other age groups) and people living in very rural communities (relative to urban and rural communities).
- Despite this, some of those at greatest risk of skin disease, including school-age children and males, remained disproportionately under-represented amongst those who had sought care; and around one in twenty individuals with a serious skin disease still incurred catastrophic out-of-pocket costs.

*What do these findings mean?:*

- Integrating health services for skin diseases into routine primary health care can successfully and substantially increase access to care, providing an effective strategy to achieve this urgent global health policy objective.
- However, integration with primary health care alone was not sufficient to ensure fair and equitable access, as some members of society continued to experience barriers to care for skin diseases.
- Beyond skin diseases, these findings can inform how routine healthcare for other public-health priorities can be better integrated into primary health care; but also the importance of identifying and targeting groups who may remain underserved by such approaches.

## BACKGROUND

Skin disease is a major contributor to morbidity, stigma, reduced quality of life and economic hardship worldwide [1–3]. In 2021, skin diseases accounted for an estimated 44.8 million disability-adjusted life-years (DALYs), ranking seventh among causes of global disability [4]. Many skin conditions are amenable to timely diagnosis and effective treatment [5, 6] and are among the most common reasons for primary care consultation [7, 8]. Yet despite their high burden and treatability, they have historically received limited attention within global health policy and health systems strengthening efforts.

Since 2022, the World Health Organization has recommended integrating services for skin-presenting neglected tropical diseases (skin NTDs) including Buruli ulcer (BU), leprosy and yaws, to accelerate progress towards control and elimination targets [9]. In 2025, the World Health Assembly recognised skin diseases as a global health priority, urging Member States to strengthen care within primary health systems and advance universal health coverage [10]. This growing policy momentum presents an important opportunity, yet robust evidence on how to operationalise comprehensive and equitable integration of skin disease services within routine primary care remains limited [11, 12].

In many low and middle-income countries, skin disease services are constrained by limited training, inconsistent access to essential medicines and diagnostics, fragmented care pathways, and out-of-pocket costs that deter timely care-seeking [13]. While early skin NTD initiatives have focused on joint case detection [14–16] and training [17, 18], fewer studies have evaluated comprehensive integration within routine primary care or assessed whether such approaches reduce financial barriers to care [19–21]. Addressing these gaps are critical: skin NTDs are among the most stigmatising skin conditions in these settings [15,16], and often reflect broader health system weaknesses - misdiagnosis, delayed treatment, and avoidable complications - that also affect people with far more common skin diseases.

In partnership with district health authorities, we designed and implemented an integrated and decentralised skin health intervention over 11 months across all 17 public health facilities in a rural district of Ghana. Formative research conducted by our team found that whilst skin NTDs were present, communities were more commonly affected by untreated infections, inflammatory conditions and wounds, and that anticipated out-of-pocket costs discouraged facility-based care-seeking [22]. In response, we co-developed a context-driven intervention that broadened scope beyond skin NTDs, decentralised diagnosis and treatment to all health facilities, and strengthened the availability of diagnostics, medicines, and dressings, to improve clinical capacity while reducing financial barriers to care [23].

We aimed to evaluate whether this integrated, decentralised model increased uptake of primary care for skin diseases, improved equity of access relative to population-level disease burden, and reduced patient treatment costs. To do so, we analysed routine health facility records, data from a population-based dermatological survey, patient interviews, and provider resource assessments.

## METHODS

### Setting

This study was conducted in Atwima Mponua (estimated population 155,254 [24]), a rural district in the southwest of Ghana (Supplementary Figure 1). Primary health services are delivered through 17 public health facilities (six Community Health Planning and Services (CHPS) compounds [25], ten health centres, and one district hospital), overseen by the District Health Directorate (DHD) through its District Health Management Team (DHMT). The district was purposively selected because no previous interventions targeting skin NTDs or any other skin diseases had been implemented. Despite Ghana’s National Health Insurance Scheme (NHIS) coverage for “skin diseases and ulcers”, our formative research identified persistent out-of-pocket payments and systemic constraints, including limited provider capacity and inconsistent availability of diagnostics and essential medicines [22]. These barriers contributed to delayed care seeking and care seeking outside the district.

### Intervention

The intervention was implemented between October 2023 and September 2024. It was developed through a series of co-creation workshops with people affected by BU, primary health workers, members of the DHMT, and national skin NTD programme managers [23]. The final intervention was conceptualised as a decentralised and integrated strategy for the care of any skin disease, in which all primary health facilities would be equipped to provide diagnosis, treatment, and management for common skin conditions, wounds, and skin NTDs. In brief, the intervention comprised three overarching activities: (1) training in the assessment and management of skin conditions for primary healthcare workers; (2) improved availability of diagnostics, medicines and dressing materials at health facilities for skin conditions, including for home-based management, and (3) community engagement to increase awareness of and promote care-seeking for skin diseases through public health facilities.

Implementation of the intervention was led by health facility staff with the support of the research team, who facilitated a five-day training-of-trainers for district staff and facility leads, followed by cascade trainings to primary health care workers. Training encompassed diagnosis and treatment of skin diseases (including skin NTDs and complex wounds), reporting tools, supply management, and community engagement. All facilities received Information, Education and Communication (IEC) materials, job-aids and patient tracking forms for complex wounds developed by the research team as part of the intervention, alongside routine reporting forms for skin NTDs. Through the district supply-chain mechanism, the research team supplied each facility with selected essential medicines and consumables necessary for diagnosis and treatment of skin diseases to be made available for all people seeking care, irrespective of NHIS enrolment (supplementary materials 1). WhatsApp groups were established between facility-level health workers, district staff, and the study team to support diagnosis and supply chain management. This support was supplemented by supervisory visits conducted by a research team clinician. Building on the formative research community engagement activities, we trained community-based surveillance volunteers (CBSVs) and school health education officers (SHEPs) to promote awareness of skin diseases through health education sessions, and to conduct community-based referral of patients with skin disease using standardized forms.

### Study Design and procedures

We evaluated the intervention using a before-and-after (pre-post) design, drawing on multiple data sources: routine health facility records; standardised patient post-care questionnaires, summaries of health facility resource use, project administration costs, and an endline cross-sectional dermatological survey of 101 randomly selected communities. Trained and supervised research assistants collected data using study-specific case reporting forms in Open Data Kit (ODK) [26].

### Health facility record review

To assess changes in primary care utilisation for skin diseases, all health facilities were requested to make routine outpatient records available to the study team for monthly review throughout the intervention period. At the district hospital, two independent study clinicians identified skin disease cases from the Lightwave Health Information Management System (LHIMS) [27] using ICD-10 diagnostic codes [28]. For all other health facilities, study team members manually screened paper-based outpatient registers for skin-related diagnoses using a predefined keyword list based on the content of the intervention training, with additional entries suggestive of skin diseases reviewed and verified by a study clinician. We additionally reviewed secondary records of care for skin disease (wound trackers for complex wounds and reporting forms for skin NTDs) at all health facilities. We retrospectively extracted pre-intervention records (January to October 2023) as the comparison (pre-intervention) period for all health facilities except for the district hospital, where data were not available.

### Patient questionnaire

To evaluate the cost of receiving care through the intervention, the research team administered a patient post-care questionnaire (PPCQ) to all patients diagnosed with BU, yaws or leprosy and to a random sample of patients who had received wound care (up to 5 per facility per month), prioritising those with complex wounds requiring multiple appointments. Sampled participants were traced using location information recorded in the health facility record and were interviewed up to three times. Proxy responses from caregivers were used for participants aged under 11 years. The PPCQ captured socio-demographic data and costs incurred to seek care for their skin conditions (including medicines, wound dressings, consultations, travel, and productivity losses) at public health facilities and other providers (e.g. traditional healers, drug sellers, facilities outside the study district), before and after their diagnosis.

### Provider resource survey

To assess resource quantities and unit costs (medical and non-medical equipment, human resources, and building spaces), the research team conducted a structured survey at all health facilities in October 2024. This comprised a questionnaire administered to facility leads (and additional facility staff if relevant), and observations of resources available at the health facility. Quantities and unit costs of resources provided by the study team to health facilities for implementation of the intervention were extracted from project records.

### Cross-sectional survey

To estimate the underlying prevalence of skin disease in the study district, a cross-sectional community survey using a two-stage screen-and-verify approach was conducted from August to September 2024. The sample size was calculated for a single proportion under varying assumptions of the prevalence of any skin condition (1 in 100 to 5 in 10,000) and intra-cluster correlation (0.002 to 0.05) at 95% confidence, 0.5 fixed relative precision, 80% participation, and a finite population of 200,000. A prevalence of 0.5 to 1.5 per 1000 individuals (for rare skin diseases) and ICC of 0.005 yielded a target sample size of 50,224 individuals in 101 clusters. All 183 communities in the district were eligible for inclusion in the survey, grouped into 175 primary sampling units (PSUs) of 500 to 1500 residents by pairing smaller and subdividing larger communities. A simple random sample of 101 PSUs was then selected for the survey.

The survey was conducted between August 6 and September 21, 2024, using compact segment sampling [29]. In each PSU, a survey enumerator and community volunteer mapped the PSU boundary and estimated the total number of households. PSUs with <200 households were surveyed entirely; larger PSUs (≥200 households) were divided into segments of approximately 50 households using physical landmarks, and two segments were randomly selected to be surveyed. In stage one of the survey, a household questionnaire (including a roster of residents, assets, and access to water and sanitation facilities) was administered to a consenting adult (≥18 years). All household members present were shown images of skin diseases [17] and asked to report if anyone in the household (including those temporarily absent) had a similar condition. In stage two of the survey, individuals reported to have a skin disease were assessed at a household visit by a study clinician not involved in the delivery of the intervention. The clinician conducted a full clinical assessment (history taking, examination, and diagnosis), including the collection of samples for point-of-care or molecular testing as appropriate. At each survey stage, households were revisited up to three times if participant(s) were absent or consent was not possible.

### Outcomes

Co-primary outcomes of the study are the incidence (as determined from health facility records) and prevalence (as determined by the survey) of (1) any skin disease, and four specific categories of skin disease: (2) complex wounds, (3) inflammatory dermatoses, (4) dermatophytosis and (5) scabies. The incidence and prevalence of other skin NTDs (BU, leprosy and yaws) are additionally reported as secondary outcomes. Any skin disease was defined as any specific dermatosis, type of lesion or other relevant description recorded in the ‘final diagnosis’ field (for health facility records); or as any active lesion as diagnosed by a study clinician (during stage two of the cross-sectional survey). During the review of health facility records, a complex wound was defined as any wound recorded at two or more consultations; for the cross-sectional survey, wounds were categorised as traumatic or non-traumatic based on patient reporting. Inflammatory dermatosis was defined as dermatitis, pruritus or any other relevant ICD-10 code. Dermatophytosis was defined as any superficial fungal infection (e.g. tinea corporis, tinea capitis, tinea pedis). Scabies was diagnosed using the clinical criteria (Level B) of the International Alliance for the Control of Scabies Consensus Criteria [30]. Diagnoses of BU, leprosy, or yaws were defined based on internationally recognised diagnostic criteria: BU by IS2404 quantitative PCR [31], leprosy by expert clinical examination (with all potential cases confirmed by a DCO) [32], and yaws by point-of-care serological testing using the Chembio DPP Screen and Confirm test [33].

### Analysis

#### Epidemiological analysis

Incidence rates were calculated as the number of cases for each co-primary outcome divided by person-time at risk (PTAR) by health facility, aggregated to estimate district-level incidence. PTAR was derived from 2024 population catchment estimates for each health facility, excluding any months for which facility health records were missing. To permit stratified analysis, age and sex-specific PTAR was calculated using publicly available census data [34]; and location-specific (very rural, rural, or urban community based on classifications provided by the DHD) PTAR was calculated from DHD population estimates for each community, scaled to match the overall (health-facility aggregated) population estimate. We estimated incidence rate ratios (IRR) by comparing the (i) intervention *versus* pre-intervention periods for CHPS compounds and health centres combined, and (ii) CHPS compounds and health centres *versus* the district hospital, both during the intervention period. Crude IRR were estimated using univariable Poisson regression and a multivariable Poisson model fitted adjusting for sex, age group, and community location.

Prevalence of co-primary outcomes were calculated as the number of cases diagnosed during stage 2 of the cross-sectional survey, divided by the total number of people enumerated during stage 1. We summarised prevalence data descriptively and estimated intra-cluster correlation coefficients (ICCs) using intercept-only mixed-effects logistic regression models. Factors associated with any skin disease were assessed using mixed-effects logistic regression with random intercepts for cluster and household. Unadjusted odds ratios were estimated for each candidate variable. For the final multivariable model, sex, age group, community location, and socioeconomic status (constructed from reported ownership of household assets using principal component analysis [35]) were included *a priori*, with selection of additional candidate variables informed by Bayesian Information Criterion (BIC). Demographic differences between survey and health facility cases were assessed using the X² test for homogeneity.

#### Economic analysis

We estimated both financial costs (expenditure) and economic costs (opportunity costs regardless of expenditure) to both patients and providers.

Patient costs comprised out-of-pocket (OOP) payments made by or on behalf of the patient in seeking care for their skin disease, and patients’ productivity losses due to the skin disease. OOP costs comprised payments made both before and during the intervention period, at both intervention and non-intervention facilities. Productivity losses were only collected for the intervention period and valued using the human capital approach by multiplying the number of working days missed by the daily minimum wage (GHS18.15=USD1.27). To assess the burden of OOP, we calculated the proportion of patients who incurred catastrophic expenditure, defined as >10% and >25% of annual household spending [36], estimated using average annual district household spending (GHC15,474.6=USD1,082.8) from the Ghana Living Standards Survey [37]. Patient costs are presented by component (medical, non-medical, or productivity losses) and stratified by age, sex, insurance status, number of facility visits, and occupation. Multiple linear regression was used to estimate the association between the financial cost per patient during the intervention period and patient characteristics. A semi-log model was estimated, with the cost variable log-transformed to reduce bias from skewness.

Provider costs included all recurrent and capital resources used during the intervention period to provide patient care for skin diseases. Health worker time spent on the care of skin diseases was estimated based on the average time per patient with skin disease and the total number of patients with skin diseases. The value of capital items (resources with a useful life >1 year) were annualised using straight-line depreciation. For resources shared between skin disease and other (non-skin disease) care, we assumed equal average use per visit. Resources were valued using 2024 unit prices from study records or market prices. Health worker salaries were estimated using the Single Spine Pay Policy. Provider costs are presented by health facility (total and per patient) and disaggregated by intervention activity and resource category.

### Ethical Approval

The study was approved by the Ghana Health Services Ethics Committee (#010/03/23); Institutional Review Board of Noguchi Memorial Institute for Medical Research (#FWA00001824); Committee of Human Research, Publication and Ethics at Kwame Nkrumah University of Science and Technology (#CHRPE/AP/153/23); and Interventions Research Ethics Committee at the London School of Hygiene and Tropical Medicine (#28445/RR/30423).

## RESULTS

### Incidence of skin disease

Health records were obtained from all 17 study health facilities for the full intervention period (November 2023 to September 2024; 187 facility-months). For the pre-intervention period (January to October 2023), health records were complete for 13 health facilities (130 facility-months), partial for three health facilities (22 facility-months), and not available for one health facility (district hospital). During the intervention period, 10,754 cases of skin disease were recorded across all health facilities, giving a district-level incidence of 74.7 per 1,000 PYAR (95% CI: 73.3–76.1). Most cases of skin disease were scabies (n=1,416), inflammatory dermatoses (1,394), dermatophytosis (1,234), or complex wounds (1,101); other skin NTDs were rare (Buruli ulcer, n=1; leprosy, n=2; yaws, n=23) (Supplementary Table 1).

At CHPS facilities and health centres, the incidence of any skin disease increased during the intervention period from 30.8 to 61.7 cases per 1,000 PYAR (aIRR=2.0, 95% CI 1.92–2.09) (Table 1 and Figure 1). Substantial increases were observed for scabies, dermatophytosis, and complex wounds, with scabies increasing most (aIRR=83.2; 95% CI 48.2–143.8). In contrast the incidence of inflammatory dermatoses declined (aIRR=0.54, 95% CI 0.50–0.62). Incidence and rate increase of any skin disease were similar by sex during the intervention period (female IR=62.5 per 1000 PYAR, 95% CI 57.8–67.6; female aIRR=2.02, 95% CI 1.90–2.15). By age group, incidence of any skin disease was greatest during the intervention period among pre-school age children (PSAC) (IR=147.2 per 1000 PYAR, 95% CI 133.2–162.7). While relative increases in skin disease occurred in all age groups, they were greatest among school-age children (SAC) (aIRR=2.78, 95% CI 2.52–3.07), as compared to PSAC or adults.

**Table 1:** Incidence of skin diseases in Atwima Mponua district; November 2023–September 2024 (intervention period) versus January–October 2023 (pre-intervention period)

|  | Intervention period *versus* pre-intervention period |  |  |  |
| --- | --- | --- | --- | --- |
|  | Incidence rate (95% CI)<br>CHPS facility and health centres |  | Crude IRR*<br>(95% CI) | Adjusted IRR†<br>(95% CI) |
|  | Pre-intervention | Intervention |  |  |
| Any skin disease | 30.8 (29.8, 31.9) | 61.7 (58.4, 65.1) | 2.00 (1.92, 2.09) | 2.00 (1.92, 2.09) |
| **By specific skin disease:** |  |  |  |  |
| Complex wound‡ | 0.2 (0.2, 0.3) | 4.0 (2.2, 7.0) | 17.06 (11.32, 25.71) | 16.61 (10.71, 25.75) |
| Inflammatory dermatoses§ | 9.4 (8.8, 10.0) | 5.1 (4.5, 5.7) | 0.54 (0.48, 0.60) | 0.55 (0.50, 0.62) |
| Ringworm | 1.0 (0.8, 1.2) | 9.0 (6.8, 11.9) | 9.11 (7.44, 11.17) | 8.83 (7.13, 10.92) |
| Scabies | 0.1 (0.1, 0.2) | 11.6 (5.6, 23.7) | 79.86 (48.01, 132.82) | 83.24 (48.2, 143.76) |
| Any other skin disease | 20.0 (19.2, 20.9) | 32.1 (30.0, 34.4) | 1.60 (1.52, 1.69) | 1.61 (1.52, 1.70) |
| **By patient characteristic:** |  |  |  |  |
| **Sex:** |  |  |  |  |
| Male | 31.1 (29.6, 32.6) | 60.3 (56.0, 65.1) | 1.94 (1.83, 2.06) | 1.98 (1.86, 2.11) |
| Female | 30.4 (28.9, 31.9) | 62.5 (57.8, 67.6) | 2.06 (1.94, 2.19) | 2.02 (1.90, 2.15) |
| **Age group:** |  |  |  |  |
| PSAC (0-4 years) | 64.0 (60.0, 68.3) | 147.2 (133.2, 162.7) | 2.30 (2.13, 2.48) | 2.28 (2.10, 2.47) |
| SAC (5-17 years) | 18.2 (16.8, 19.8) | 49.3 (43.6, 55.8) | 2.70 (2.46, 2.97) | 2.78 (2.52, 3.07) |
| Young adult (18-39 years) | 26.1 (24.4, 27.9) | 39.9 (35.9, 44.3) | 1.52 (1.41, 1.66) | 1.54 (1.42, 1.68) |
| Older adult (≥40 years) | 34.1 (31.7, 36.6) | 54.4 (48.6, 60.9) | 1.60 (1.46, 1.74) | 1.63 (1.48, 1.78) |
| **Place of residence:** |  |  |  |  |
| Very rural | 14.0 (12.6, 15.6) | 39.0 (33.2, 45.8) | 2.79 (2.47, 3.15) | 2.76 (2.45, 3.12) |
| Rural | 27.7 (26.3, 29.2) | 51.7 (47.6, 56.1) | 1.87 (1.75, 1.99) | 1.85 (1.73, 1.97) |
| Urban | 39.4 (37.1, 41.7) | 77.6 (70.8, 85.1) | 1.97 (1.84, 2.12) | 1.97 (1.83, 2.10) |
| **By facility type:** |  |  |  |  |
| Level 1 (CHPS) | 14.0 (12.9, 15.2) | 49.0 (43.5, 55.3) | 3.50 (3.20, 3.83) | - |
| Level 2 (Health centre) | 42.4 (40.8, 44.1) | 69.9 (65.7, 74.3) | 1.65 (1.57, 1.73) |  |

IRR = incidence rate ratio; CI = confidence interval, PSAC = Pre-school age children, SAC = School age children.
Incidence is expressed as cases per 1,000 resident population per year. Incidence from health records of 16 government health facilities within Atwima Mponua district are reported: level 1 (Community Health Planning and Services (CHPS) facility, n=6) and level 2 (Health centre, n=10). \* Total of 10 650 observations. Missing data for sex (n=40); age group (n=130); place of residence (n=1 037). † Total of 9 472 observations, mutually adjusted for patient sex, age group, and place of residence. ‡: Defined as a recorded diagnosis of [Dermatitis] or [Pruritus]. §: Defined as an individual with two or more records of [Wound] or [Ulcer] within a two week period (pre-intervention period); or an individual registered on a 'wound tracker' or recorded in OPD register with two or more records of [Wound] or [Ulcer] within a two week period (intervention period).

**Figure 1:**
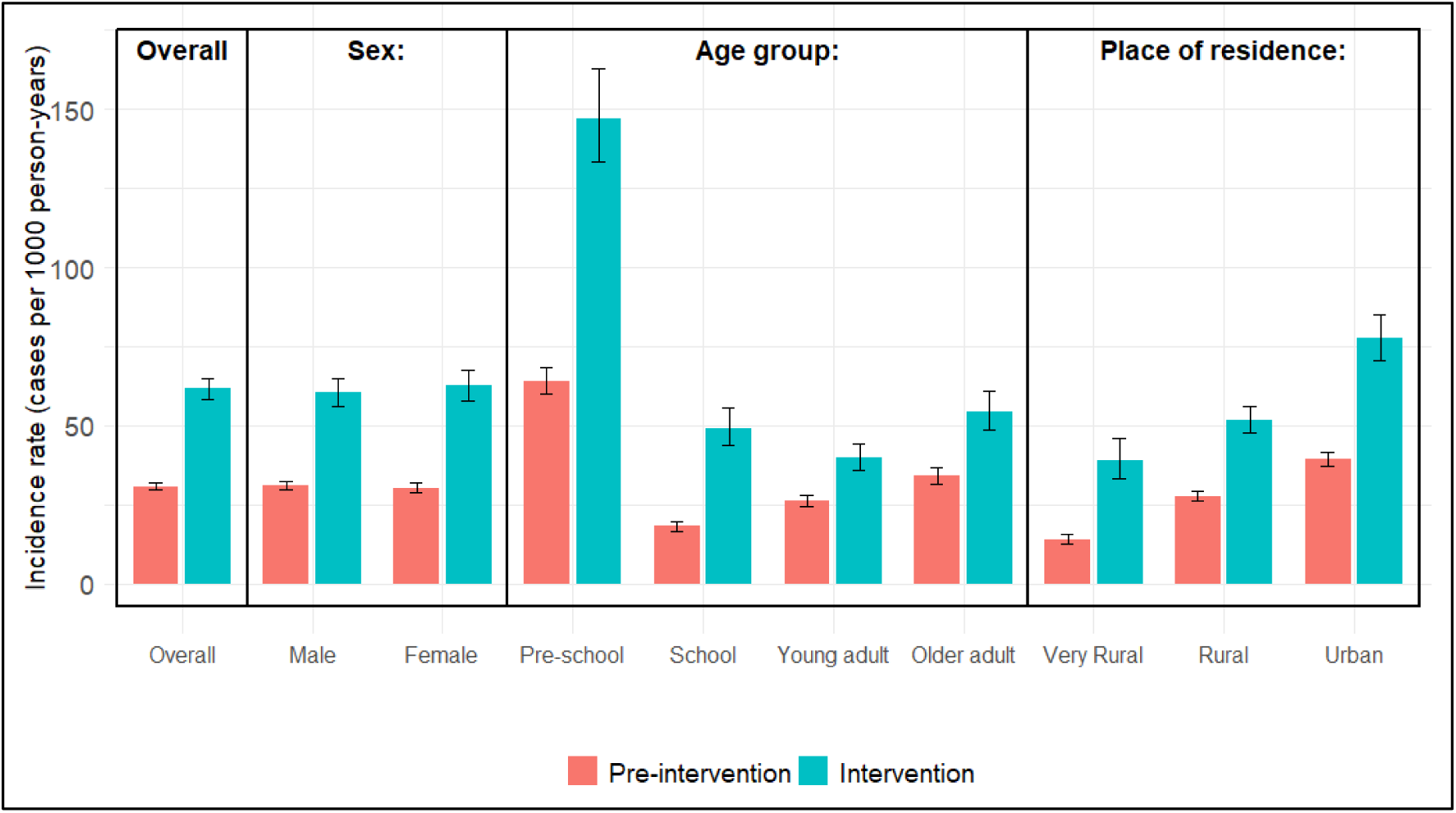
Incidence rates of any diagnosed skin disease; pre-intervention period (January–October 2023) versus intervention period (November 2023–September 2024)

During the intervention period, incidence of skin disease was lowest in very rural communities (IR=39.0 per 1,000 PYAR; 95% CI 33.2–45.8) and highest in urban communities (IR=77.6; 95% CI 70.8–85.1). However, the relative increase in skin disease was greatest in very rural communities (aIRR=2.76; 95% CI 2.45–3.12). This pattern was reflected at the health facility level. CHPS facilities, which primarily serve very rural populations, showed a greater increase in cases (aIRR = 3.50; 95% CI 3.20–3.83) compared to health centres. Nevertheless, the overall incidence of skin disease during the intervention period remained lower at CHPS facilities and health centres than at the district hospital (aIRR=0.42; 95% CI 0.41–0.44) (Supplementary Table 2). This pattern was consistent across sex and age groups, though it varied substantially by specific skin disease.

### Prevalence of skin disease

A total of 23,398 structures were identified in 101 clusters, of which 13,417 (57.3%) were residential (Figure 2). Among these, 11,522 residential structures (85.9%) had at least one adult present, of whom 10,970 (95.2%) consented to participate in the survey. A total of 42,801 individuals were enumerated, of whom 4,250 (9.9%) reported a skin disease. Clinical verification was attempted for 4,175 individuals (98.2%), of whom3,507 (84.0%) were available. After excluding 1,080 individuals who were not eligible for assessment (no current skin disease, n=1062; or did not consent, n=18), and the further inclusion of 55 individuals who reported skin disease during verification, 2,482 individuals were assessed for skin disease.

**Figure 2:**
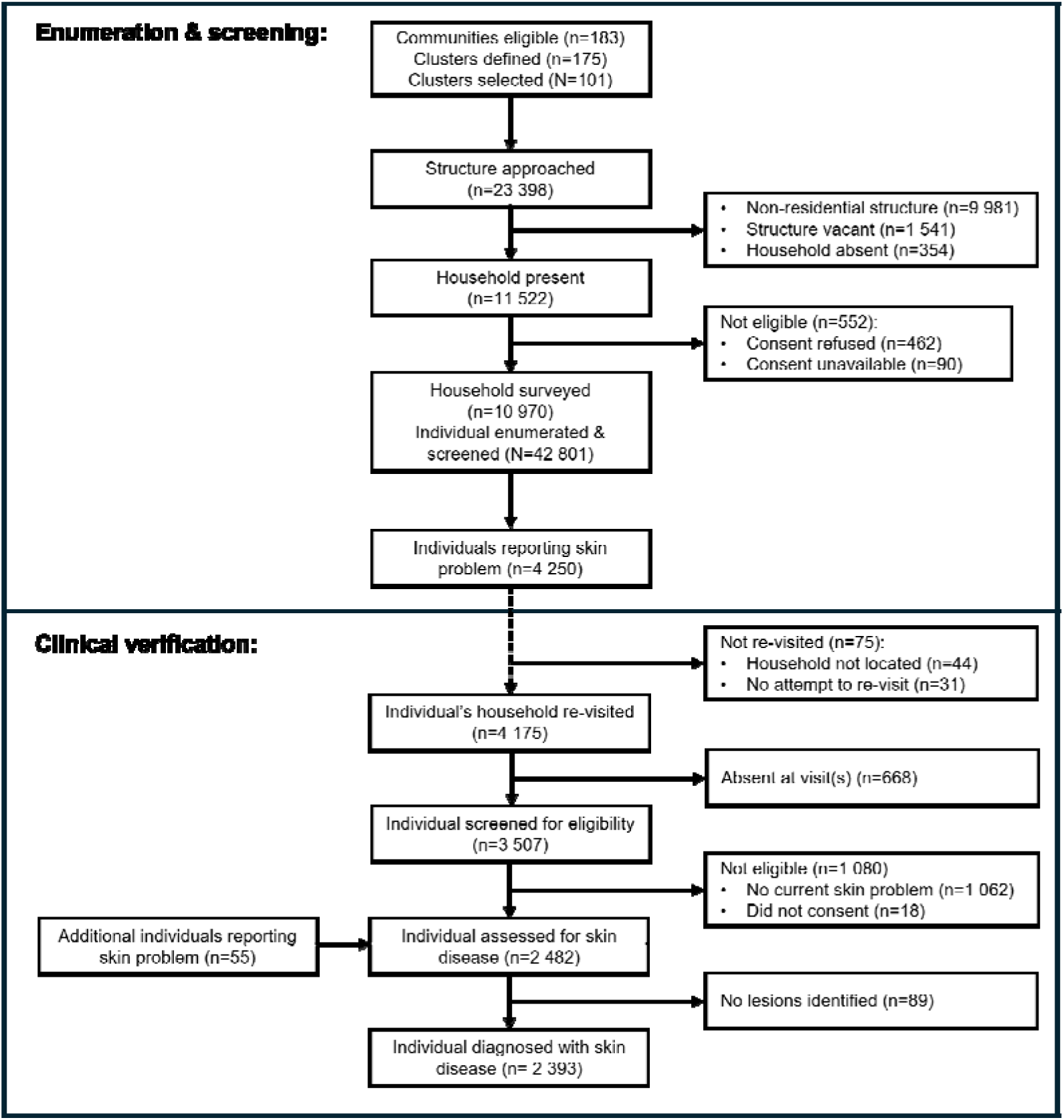
**Cross-sectional survey flowchart**

A total of 2,393 individuals were diagnosed with a skin disease, corresponding to a prevalence of 559 per 10,000 persons (95% CI 538–581). A greater proportion of cases were amongst males (53.0%) and children or young adults (median age=10; IQR:5–24) (Supplementary Table 3). The most prevalent skin diseases were dermatophytosis (255 per 10,000 persons, 95% CI 240–271), scabies (133 per 10,000 persons, 95% CI 123–145), and traumatic wounds (45 per 10,000 persons, 95% CI 39–52). Skin NTDs other than scabies were rare (Buruli ulcer: n=1; leprosy: n=5; yaws: n=8), with Buruli ulcer and leprosy observed only amongst adults, and yaws primarily amongst children (median age: 10.5; IQR: 8.5–13). Strong clustering was observed for yaws (ICC=0.80) and scabies (ICC=0.31), compared to for the presence of any skin disease (ICC=0.10).

In multivariate analysis, odds of skin disease were significantly higher amongst males (aOR=1.27, 95% CI 1.15–1.40) and PSAC (aOR=1.72, 95% CI 1.11–2.67) as compared to SAC and adults (Table 2). Lower odds of skin disease were observed among individuals with lower secondary education (aOR=0.52, 95% CI 0.41–0.66), or a manual (aOR=0.52, 95% CI 0.34–0.79) or non-manual occupation (aOR=0.66, 95% CI 0.44–0.49) (relative to those with lower levels of education or no occupation). Odds of skin disease were higher when the head of household had primary (aOR=1.34, 95% CI 1.09–1.66) or higher levels (aOR=1.16, 95% CI 1.00–1.34) of education. We observed no evidence of association between skin disease and household socioeconomic status (SES), or having recently moved into the district. At the community level, odds of skin disease were significantly higher in very rural communities (aOR=1.71, 95% CI 1.13–2.56) as compared to urban or rural communities, although we observed no association with the distance of the household from a health facility, or the presence of an active CBSV in the community.

**Table 2:**
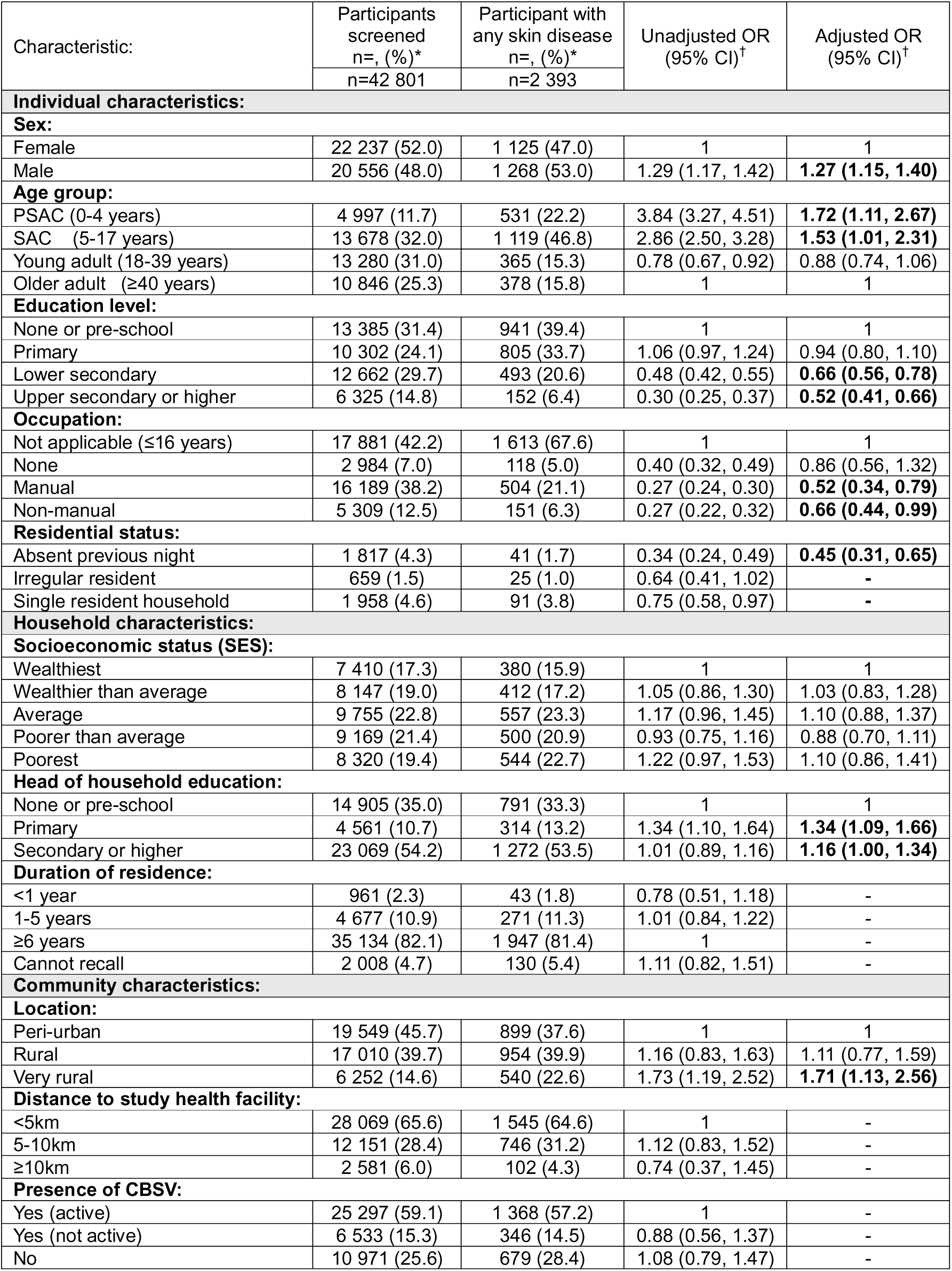
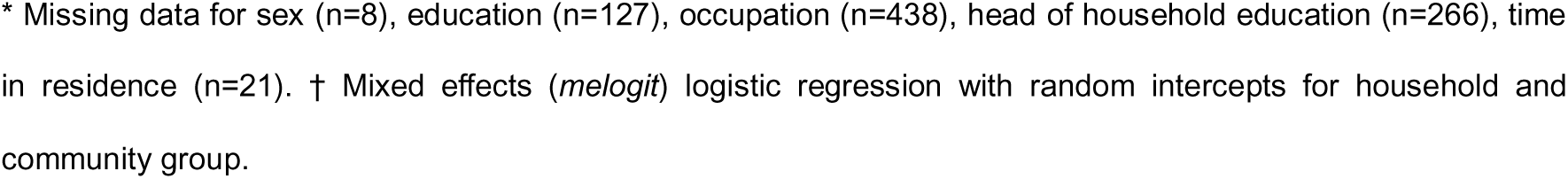
Factors associated with presence of any skin disease in Atwima Mponua district, amongst cross-sectional survey participants (July–September 2024)

| Characteristic: | Participants screened<br>n=, (%)<sup>*</sup> | Participant with any skin disease<br>n=, (%)<sup>*</sup> | Unadjusted OR<br>(95% CI)<sup>†</sup> | Adjusted OR<br>(95% CI)<sup>†</sup> |
| --- | --- | --- | --- | --- |
| **Individual characteristics:** |  |  |  |  |
| **Sex:** |  |  |  |  |
| Female | 22 237 (52.0) | 1 125 (47.0) | 1 | 1 |
| Male | 20 556 (48.0) | 1 268 (53.0) | 1.29 (1.17, 1.42) | **1.27 (1.15, 1.40)** |
| **Age group:** |  |  |  |  |
| PSAC (0-4 years) | 4 997 (11.7) | 531 (22.2) | 3.84 (3.27, 4.51) | **1.72 (1.11, 2.67)** |
| SAC (5-17 years) | 13 678 (32.0) | 1 119 (46.8) | 2.86 (2.50, 3.28) | **1.53 (1.01, 2.31)** |
| Young adult (18-39 years) | 13 280 (31.0) | 365 (15.3) | 0.78 (0.67, 0.92) | 0.88 (0.74, 1.06) |
| Older adult (≥40 years) | 10 846 (25.3) | 378 (15.8) | 1 | 1 |
| **Education level:** |  |  |  |  |
| None or pre-school | 13 385 (31.4) | 941 (39.4) | 1 | 1 |
| Primary | 10 302 (24.1) | 805 (33.7) | 1.06 (0.97, 1.24) | 0.94 (0.80, 1.10) |
| Lower secondary | 12 662 (29.7) | 493 (20.6) | 0.48 (0.42, 0.55) | **0.66 (0.56, 0.78)** |
| Upper secondary or higher | 6 325 (14.8) | 152 (6.4) | 0.30 (0.25, 0.37) | **0.52 (0.41, 0.66)** |
| **Occupation:** |  |  |  |  |
| Not applicable (≤16 years) | 17 881 (42.2) | 1 613 (67.6) | 1 | 1 |
| None | 2 984 (7.0) | 118 (5.0) | 0.40 (0.32, 0.49) | 0.86 (0.56, 1.32) |
| Manual | 16 189 (38.2) | 504 (21.1) | 0.27 (0.24, 0.30) | **0.52 (0.34, 0.79)** |
| Non-manual | 5 309 (12.5) | 151 (6.3) | 0.27 (0.22, 0.32) | **0.66 (0.44, 0.99)** |
| **Residential status:** |  |  |  |  |
| Absent previous night | 1 817 (4.3) | 41 (1.7) | 0.34 (0.24, 0.49) | **0.45 (0.31, 0.65)** |
| Irregular resident | 659 (1.5) | 25 (1.0) | 0.64 (0.41, 1.02) | - |
| Single resident household | 1 958 (4.6) | 91 (3.8) | 0.75 (0.58, 0.97) | - |
| **Household characteristics:** |  |  |  |  |
| **Socioeconomic status (SES):** |  |  |  |  |
| Wealthiest | 7 410 (17.3) | 380 (15.9) | 1 | 1 |
| Wealthier than average | 8 147 (19.0) | 412 (17.2) | 1.05 (0.86, 1.30) | 1.03 (0.83, 1.28) |
| Average | 9 755 (22.8) | 557 (23.3) | 1.17 (0.96, 1.45) | 1.10 (0.88, 1.37) |
| Poorer than average | 9 169 (21.4) | 500 (20.9) | 0.93 (0.75, 1.16) | 0.88 (0.70, 1.11) |
| Poorest | 8 320 (19.4) | 544 (22.7) | 1.22 (0.97, 1.53) | 1.10 (0.86, 1.41) |
| **Head of household education:** |  |  |  |  |
| None or pre-school | 14 905 (35.0) | 791 (33.3) | 1 | 1 |
| Primary | 4 561 (10.7) | 314 (13.2) | 1.34 (1.10, 1.64) | **1.34 (1.09, 1.66)** |
| Secondary or higher | 23 069 (54.2) | 1 272 (53.5) | 1.01 (0.89, 1.16) | **1.16 (1.00, 1.34)** |
| **Duration of residence:** |  |  |  |  |
| <1 year | 961 (2.3) | 43 (1.8) | 0.78 (0.51, 1.18) | - |
| 1-5 years | 4 677 (10.9) | 271 (11.3) | 1.01 (0.84, 1.22) | - |
| ≥6 years | 35 134 (82.1) | 1 947 (81.4) | 1 | - |
| Cannot recall | 2 008 (4.7) | 130 (5.4) | 1.11 (0.82, 1.51) | - |
| **Community characteristics:** |  |  |  |  |
| **Location:** |  |  |  |  |
| Peri-urban | 19 549 (45.7) | 899 (37.6) | 1 | 1 |
| Rural | 17 010 (39.7) | 954 (39.9) | 1.16 (0.83, 1.63) | 1.11 (0.77, 1.59) |
| Very rural | 6 252 (14.6) | 540 (22.6) | 1.73 (1.19, 2.52) | **1.71 (1.13, 2.56)** |
| **Distance to study health facility:** |  |  |  |  |
| <5km | 28 069 (65.6) | 1 545 (64.6) | 1 | - |
| 5-10km | 12 151 (28.4) | 746 (31.2) | 1.12 (0.83, 1.52) | - |
| ≥10km | 2 581 (6.0) | 102 (4.3) | 0.74 (0.37, 1.45) | - |
| **Presence of CBSV:** |  |  |  |  |
| Yes (active) | 25 297 (59.1) | 1 368 (57.2) | 1 | - |
| Yes (not active) | 6 533 (15.3) | 346 (14.5) | 0.88 (0.56, 1.37) | - |
| No | 10 971 (25.6) | 679 (28.4) | 1.08 (0.79, 1.47) | - |

\* Missing data for sex (n=8), education (n=127), occupation (n=438), head of household education (n=266), time in residence (n=21). † Mixed effects (*me**logit*) logistic regression with random intercepts for household and community group.

Overall, 30.2% of survey participants with skin disease reported visiting a health facility for their condition (Supplementary Table 3). This proportion was consistently low for common skin diseases, including dermatophytosis (23.2%), inflammatory dermatosis (32.4%), and scabies (35.4%). A significantly larger proportion of participants with non-traumatic wounds had visited a health facility than those with traumatic wounds (62.5% *versus* 28.4%, p<0.001). No participant with yaws (n=8) and 2 of 5 people affected by leprosy identified in the survey had visited a health facility, whereas the one participant with suspected Buruli ulcer was currently receiving care.

### Comparison between health facility records and cross-sectional survey

Demographic comparison of those who sought care during the intervention period versus those identified during the cross-sectional survey revealed significant disparities in the burden of skin disease (Table 3). Relative to the prevalence of any skin disease, males were under-represented among those with skin disease in health facility records (absolute difference in proportion= -5.3%), while females were over-represented (+5.2%; p=0.04). Similarly, we observed disparities by age group, with children substantially under-represented in health facility records (PSAC: -7.2%; SAC: -24.0%), and adult over-represented (younger adults: +8.9%; older adults: +7.7%, p<0.001). Finally, we observed a trend towards under-representation of individuals from very rural communities (-8.2%) compared to those from rural (+2.9%) and peri-urban (+5.3%) areas, though this difference was not significant after adjusting for survey design (p=0.54). When disaggregated by skin disease case type, inflammatory dermatoses (-8.9%), dermatophytosis (-31.4%), and scabies (-9.2%) comprised a significantly smaller proportion of skin diseases recorded in health facility records, as compared to the cross-sectional survey. Conversely, wounds of any class (+11.2%) and any other skin disease (+20.8%) comprised a significantly greater proportion. Finally, there was no evidence of correlation between the standardized community-level prevalence of skin conditions at the endline survey and standardized community-level incidence based on health facility records (R2 (coefficient of determination) 0.006).

**Table 3:** Comparison of prevalence of skin disease (cross-sectional survey) *versus* incidence of skin disease (health facility records) in Atwima Mponua district; July–September 2024 (cross-sectional survey) and November 2023–September 2024 (intervention period)

| Characteristic: | Survey participant with any skin disease<br>n=2 393 | Patient with any skin disease<br>n=10 754<sup>1</sup> | Difference in proportion<sup>2</sup> | Test statistic<sup>3</sup> |
| --- | --- | --- | --- | --- |
| Sex: |  |  |  |  |
| Male | 1 268 (53.0) | 5 126 (47.7) | -5.3 | 0.04 |
| Female | 1 125 (47.0) | 5 596 (52.2) | +5.2 |  |
| Age group: |  |  |  |  |
| PSAC (0-4 years) | 531 (22.2) | 3 133 (29.4) | -7.2 | <0.001 |
| SAC (5-17 years) | 1 119 (46.8) | 2 427 (22.8) | -24.0 |  |
| Young adult (18-39 years) | 365 (15.3) | 2 577 (24.2) | +8.9 |  |
| Older adult (≥40 years) | 378 (15.8) | 2 504 (23.5) | +7.7 |  |
| Location: |  |  |  |  |
| Very rural | 540 (22.6) | 1 113 (14.4) | -8.2 | 0.54 |
| Rural | 954 (39.9) | 3 310 (42.8) | +2.9 |  |
| Peri-urban | 899 (37.6) | 3 320 (42.9) | +5.3 |  |
|  | Survey participant skin disease case<br>n=2 550 | Patient with any skin disease<br>n=10 754<sup>1</sup> |  |  |
| Specific diagnosis: |  |  |  |  |
| Inflammatory dermatoses | 105 (4.1) | 1394 (13.0) | -8.9 | - |
| Ringworm | 1 094 (42.9) | 1234 (11.5) | -31.4 |  |
| Scabies | 570 (22.4) | 1416 (13.2) | -9.2 |  |
| Wound (traumatic) | 194 (7.6) | 2229 (20.7) | 11.2 |  |
| Wound (non-traumatic) | 48 (1.9) |  |  |  |
| BU, leprosy or yaws | 14 (0.5) | 26 (0.2) | -0.3 |  |
| Other skin disease | 525 (20.6) | 4455 (41.4) | 20.8 |  |

1: Missing data: sex (n=32), age group (n=113), location (n=3 011). 2: Intervention proportion (patient with any skin disease) minus survey proportion (patient with any skin disease). 3: X<sup>2</sup> test for homogeneity.

### Costs to patients with skin NTDs and complex wounds

Patients seeking care for skin NTDs and complex wounds incurred a median financial cost of GHS 90 (USD 6.5) and a mean financial cost of GHC 1041 (USD 72.80) (Supplementary Table 4). Of the total costs, 79% (GHS 826, USD 57.76) were incurred before the first visit to a study health facility during the intervention period (index visit). These pre-visit costs were primarily driven by Consultation fees (43%; GHS 340, USD 23.78) and medicines (28%; GHS 222, USD 15.52), incurred through visits to private providers, drug sellers, traditional healers or GHS health facilities outside the study district. During and after the index visit, the mean costs of consultation (GHS 29, USD 2.3) and medicines (GHS 55, USD 3.85) were substantially lower. Costs per patient were right-skewed, with most patients incurring costs lower than the mean and cost drivers varying across patients. Approximately 4% (n=16) and 0.6% (n=4) of patients with skin NTDs and complex wounds experienced catastrophic expenditure at the >10% and >25% thresholds), respectively (Figure 3).

**Figure 3:**
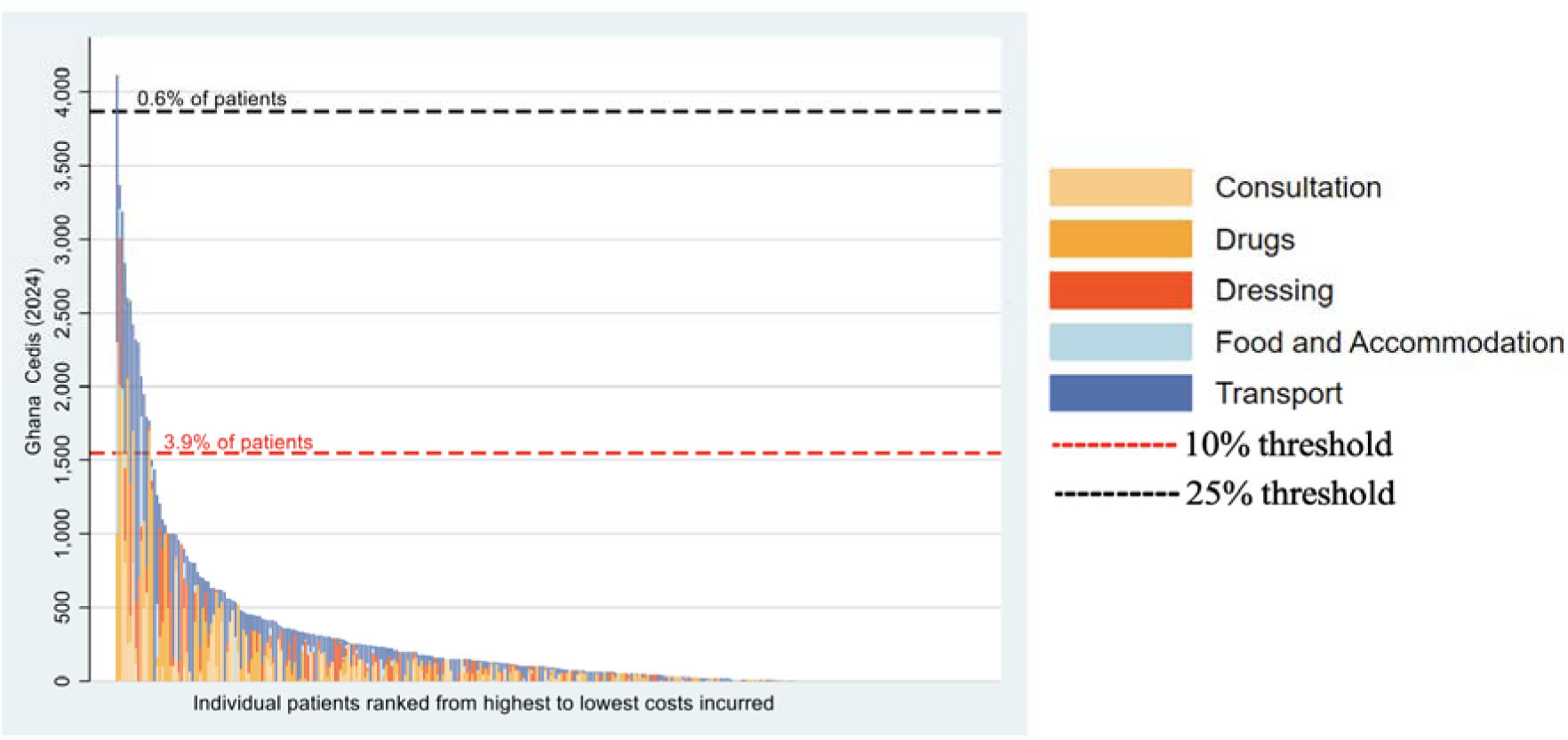
Catastrophic expenditure and individual patient costs by category Red and black dotted lines: 10% and 25%, respectively, of average household total consumption expenditure in Atwima Mponua District.

During and after the index visit, productivity losses from care-seeking cost an additional mean of GHS 373 (USD 26.07) per patient with skin NTDs or complex wounds (Supplementary Table 5). The mean economic cost of care-seeking was GHS 540 (USD 37.76), and the median was GHS 197 (USD 13.78). In multiple linear regression, mean financial costs were 142% (p<0.001) and 128% (p=0.011) higher among patients aged 18-60 and >60 years of age, respectively, relative to those aged <18 years. Relative to those making <4 visits for their skin condition, those making 4-10 visits experienced 119% (p=0.001) higher mean costs, and those making >10 visits experienced 194% (p<0.001) higher mean costs. Other patient characteristics did not have statistically significant associations with patient financial costs. NHIS members had similar mean financial costs (GHS 220, USD 15.38) to non-members (GHS 204, USD 14.27), with both groups incurring out-of-pocket costs. We did not observe evidence of mean financial cost variation across wealth quintiles.

### Costs to providers for all skin diseases

From a provider perspective, the intervention had a total cost of GHS1,539,679 (US$107,670) and mean cost of GHS 183 (USD 12.80) per patient treated, ranging from GHS 173 to 202 (USD 12.10 to 14.13) across CHPS facilities, GHS169 to 216 (USD 11.82 to 15.11) across health centres, and with a mean of GHS 177 (USD 12.38) at the district hospital (Supplementary Figure 3). Clinical care was the largest cost driver, accounting for 42% of costs, followed by training (31%), community engagement (17%), preparation (8%), and monitoring and evaluation (2%). Consumables and human resources were consistently the main drivers of provider costs across all types of health facility. Costs of clinical care, training, and community engagement (CE) activities were driven by consumables and human resources costs, while for training, transportation costs were also substantial (Supplementary Figure 4).

## DISCUSSION

This study provides evidence that integrating skin NTD services with broader skin disease care within routine primary care can substantially increase service uptake at district scale. Following implementation of a district-wide intervention, recorded incidence of skin disease doubled, with the largest relative gains among children and residents of very rural communities. However, comparison of facility and community survey data revealed persistent inequities, with many individuals - particularly men, children, and those in very rural areas - not seeking care. Although patient costs were generally low once care was accessed and catastrophic expenditure was uncommon, expenditures were high prior to presentation at intervention health facilities. Together, these findings suggest that integration can expand access, but equitable and financially protective impact requires deliberate strategies to reach underserved and higher risk populations.

Our results add to emerging evidence that integrating skin disease services into primary care can improve case detection and coverage when supported by strengthened workforce capacity and information systems, and by combining active case finding and facility-or mobile-clinic based follow up [20, 38–40]. Experience from leprosy integration demonstrates that such gains are sustained only when supply chains are reliable, referral pathways functional, and case management embedded within existing primary care workflows rather than operating as parallel vertical programmes [41]. Extending this evidence, we demonstrate that comprehensive management of skin NTDs, complex wounds, and common high-burden skin conditions can be embedded within routine district primary care systems at scale – provided that health workers are trained and supported to manage both common and rare conditions, and that essential diagnostics and medicines are consistently available. The intervention sought to address gaps between NHIS coverage and practical access by improving facility-level availability of essential medicines and dressings. Although this reduced patient costs once patients presented to intervention facilities, stock gaps persisted at times, and patients incurred out-of-pocket costs when NHIS-covered commodities were unavailable. These findings highlight that integration requires not only training and decentralisation, but resilient supply systems capable of sustaining reliable access to insured services.

Consistent with previous studies in Africa [42, 43], we found a high prevalence of bacterial, and fungal skin infections and traumatic wounds - conditions frequently overlooked by interventions addressing rarer conditions such as skin NTDs and chronic ulcers. While previous integration efforts have sought to expand access to diagnosis by training frontline health workers or linking them to specialist support through teledermatology [44, 45], few have examined which populations remains excluded. Despite substantial increases in care-seeking, school-age children and males remained underrepresented amongst facility attendees relative to their overall burden of skin disease. These disparities may reflect perceptions that common skin diseases are minor, opportunity costs associated with care-seeking, and structural barriers affecting caregivers and working-age men – patterns also described in care pathways for other acute and chronic conditions in similar settings [46, 47]. While active case detection is often used for high morbidity, low prevalence conditions such as skin NTDs, it may be impractical to apply at scale for common skin conditions. Instead, integration strategies may need to leverage existing platforms – such as school-based services and community health workers – to reduce inequities in access without overextending primary care systems [48].

This study has limitations. The non-randomised design and absence of a contemporaneous control group limit causal inference. While under-reporting of skin disease prior to the intervention is unlikely given the use of facility records for NHIS reimbursement, improvements in diagnostic specificity during the intervention may have influenced condition-specific incidence trends. Missing pre-intervention hospital health records preclude assessment of potential shifts from hospital to health centres and CHPS compounds. In the cross-sectional survey, reliance on self-report prior to clinical verification and some loss to follow-up may have contributed to underestimation of true burden. We did not conduct a full cost-effectiveness analysis. However, implementation across all 17 facilities within the district’s routine health system, with minimal external support, enhances the relevance of findings for real-world scale-up, although longer follow-up is needed to assess sustainability.

Integration of health services into primary care presents a pragmatic opportunity to address interconnected burdens of ill-health, stigma and financial hardship. Sustainable care models must prioritise both common, high-burden skin problems and rare and stigmatising NTDs, while explicitly addressing inequities in access. Taken together, our findings suggest that effective integration is likely to require sustained clinical capacity through ongoing training and mentorship, decentralised service delivery through existing district health structures, and reliable availability of diagnostics and essential medicines so that insurance entitlements are met in practice. By demonstrating both the potential and the constraints of this intervention, our study offers practical evidence to guide the incorporation of comprehensive skin health services into routine primary care in ways that advance equity and universal health coverage.

## Data Availability

Data will be made available on publication through the LSHTM Data Compass (http://datacompass.lshtm.ac.uk). Requests for release of the data will be reviewed by the relevant institutional review boards.

