## Supplementary Materials for "Effect, equity and costs of an integrated and decentralised intervention to improve access to primary care for skin diseases: a prospective before-and-after study in south-west Ghana"

Supplementary Table 1: Number of skin disease cases in Atwima Mponua district between January–October 2023 (pre-intervention period) and November 2023–September 2024 (intervention period)

| **Skin disease:** | **Number of skin disease cases:** | | |
| --- | --- | --- | --- |
|  | **Pre-intervention**  **period** | **Intervention**  **period** | |
|  | CHPS facilities & health centres^*^ | CHPS facilities & health centres^†^ | District hospital^‡^ |
| Any skin disease | 3 192 | 7 458 | 3 296 |
| **By specific skin disease:** | | | |
| Complex wound | 24 | 478 | 623 |
| Inflammatory dermatosis | 976 | 616 | 778 |
| Ringworm | 102 | 1 085 | 149 |
| Scabies | 15 | 1 398 | 18 |
| **Any other skin disease:** | | | |
| Buruli ulcer | 0 | 1 | 0 |
| Leprosy | 0 | 2 | 0 |
| Yaws (confirmed) | 0 | 5 | 0 |
| Yaws (unconfirmed) | 6 | 18 | 0 |
| Any other wound | 1 169 | 893 | 235 |
| Any other non-wound | 900 | 2 962 | 1 493 |

Notes: Buruli ulcer, leprosy and yaws (confirmed) defined as cases recorded on skin NTD01 form with confirmed (positive) diagnosis. Yaws (unconfirmed) diagnosis defined as clinical diagnosis of ‘Yaws’ recorded in consulting room register only. *: Total of 103 618 person-years at risk: 3 facilities with missing data (6, 7 and 9 months respectively) and 13 facilities with no missing data (10 months). †: Total of 120 933 person-years at risk: 16 facilities with no missing data (11 months). ‡: total of 23 070 person-years at risk: 1 facility with no missing data (11 months).

Supplementary Table 2: Incidence of skin diseases in Atwima Mponua district at district hospital *versus* CHPS facilities and health centres, within intervention period (November 2023–September 2024)

|  | **Intervention period**  (District hospital *versus* CHPS facilities and health centres) | | |
| --- | --- | --- | --- |
|  | **Incidence rate**  **(95% CI)**  District hospital | **Crude IRR**^*^  **(95% CI)** | **Adjusted IRR**^†^  **(95% CI)** |
|  | **Intervention** |  |  |
| Any skin disease | 142.9 (138.1, 147.8) | 0.43 (0.41, 0.45) | 0.42 (0.41, 0.44) |
| **By specific skin disease:** | | | |
| Complex wound^‡^ | 27.0 (25.0, 29.2) | 0.15 (0.13, 0.16) | 0.14 (0.12, 0.16) |
| Inflammatory dermatoses^§^ | 33.7 (31.4, 36.2) | 0.15 (0.14, 0.17) | 0.15 (0.13, 0.17) |
| Ringworm | 6.46 (5.50, 7.58) | 1.39 (1.17, 1.65) | 1.38 (1.16, 1.63) |
| Scabies | 0.8 (0.5, 1.2) | 14.81 (9.31, 23.59) | 14.51 (9.11, 23.10) |
| *Any other skin disease* | 74.9 (71.45, 78.52) | 0.43 (0.40, 0.45) | 0.42 (0.40, 0.45) |
| **By patient characteristic:** | | | |
| **Sex:** | | | |
| Male | 113.7 (107.8, 119.9) | 0.53 (0.50, 0.56) | 0.53 (0.49, 0.56) |
| Female | 174.1 (166.5, 182.0) | 0.36 (0.34, 0.38) | 0.35 (0.33, 0.37) |
| **Age group:** | | | |
| PSAC (0-4 years) | 213.5 (198.0, 230.2) | 0.69 (0.63, 0.75) | 0.69 (0.63, 0.75) |
| SAC (5-17 years) | 79.0 (72.8, 85.8) | 0.62 (0.57, 0.69) | 0.62 (0.57, 0.68) |
| Young adult (18-39 years) | 126.2 (118.5, 134.4) | 0.32 (0.29, 0.34) | 0.32 (0.29, 0.34) |
| Older adult (≥40 years) | 214.4 (202.0, 227.6) | 0.25 (0.23, 0.27) | 0.25 (0.23, 0.27) |
| **Place of residence:** | | | |
| Very rural | [Insufficient observations] | | - |
| Rural | 23.1 (20.4, 26.1) | 2.37 (2.08, 2.69) |  |
| Urban | 95.5 (88.87, 102.56) | 0.66 (0.60, 0.71) |  |

IRR = incidence rate ratio; CI = confidence interval, PSAC = Pre-school age children, SAC = School age children. Incidence is expressed as cases per 1,000 resident population per year. *: Total of 10 754 observations. Missing data for sex (n=32); age group (n=113); place of residence (n=3 011). †: Total of 10 619 observations, mutually adjusted for patient sex and age group only. ‡: Defined as a recorded diagnosis of [Dermatitis] or [Pruritus] at CHPS facility or health centre; or a relevant [ICD-10 code] diagnosis at district hospital. §: Defined as an individual with two or more records of [Wound] or [Ulcer] within a two week period (pre-intervention: CHPS facility or health centre); an individual registered on a ‘wound tracker’ OR an individual recorded in OPD register with two or more records of [Wound] or [Ulcer] within a two week period (intervention: CHPS facility or health centre); or an individual with two or more diagnostic or procedural codes classified as wound or ulcer related within a two week period (intervention: district hospital).

Supplementary Table 3: Prevalence of any skin disease amongst cross-sectional survey participants in Atwima Mponua district; July–September 2024

| Characteristic: | Participant with skin disease | Survey prevalence | Age (years) | Sex (male) | Known to health system* | ICC |
| --- | --- | --- | --- | --- | --- | --- |
|  | n= | /10 000 persons  (95% CI) | Median (IQR) | % | % |  |
| Any skin disease | 2 393 | 559.1 (537.5, 581.3) | 10 (5, 24) | 53.0 | 30.2 | 0.10 |
| **Non skin-NTD:** | | | | |  |  |
| Inflammatory dermatoses | 105 | 24.5 (20.1, 29.7) | 18 (12, 42) | 42.9 | 32.4 | 0.13 |
| Ringworm | 1 094 | 255.1 (240.4, 270.5) | 10 (6, 18) | 58.6 | 23.2 | 0.15 |
| Scabies | 570 | 133.2 (122.5, 144.5) | 9 (4, 21) | 49.1 | 35.4 | 0.31 |
| **Skin NTD:** |  |  |  |  |  |  |
| Buruli ulcer | 1 | 0.2 (0.01, 1.3) | 56 (-) | 0 | 100.0 | - |
| Leprosy | 5 | 1.2 (0.4, 2.7) | 39 (37, 56) | 60.0 | 60.0 | - |
| Active yaws | 8 | 1.9 (0.8, 3.7) | 10.5 (8.5, 13) | 50.0 | 0 | 0.80 |
| **Wound:** |  |  |  |  |  |  |
| Wound (traumatic) | 194 | 45.3 (39.2, 52.2) | 14 (8, 43) | 59.3 | 28.4 | 0.18 |
| Wound (non-traumatic)^†^ | 48 | 11.2 (8.3, 14.9) | 42.5 (15, 55) | 54.2 | 62.5 | 0.26 |
| **Other:** |  |  |  |  |  |  |
| Any other skin disease^‡^ | 525 | 112.7 (112.4, 133.5) | 9 (3, 24) | 46.2 | 38.7 | 0.13 |

*: Defined as those reporting *current* or *ever* receiving care from a health facility for the reported skin disease. †: Includes cases suspected for buruli ulcer (ulcerative) and suspected active yaws (ulcer). ‡: Includes cases suspected for scabies *only* but negative final diagnosis (n=1), or leprosy *only* but negative final diagnosis (n=2)

Supplementary Table 4: Mean and median financial cost of care seeking for patients with skin NTDs and complex wounds, by patient characteristics and visit time point

| Characteristic: | Before index visit | | During and after index visit | | | | | | Regression analysis | |
| --- | --- | --- | --- | --- | --- | --- | --- | --- | --- | --- |
|  |  |  | Study facility | | Non-study facility | | Combined | |  |  |
|  | Mean | Median | Mean | Median | Mean | Median | Mean | Median | Coefficient* | p-value |
| Sex: | | | | | | | | | | |
| Female | 800.35 | 150 | 155.63 | 70 | 352.4 | 162.65 | 196.03 | 76.46 | REF | - |
| Male | 849.48 | 206.33 | 189.99 | 79.09 | 262.07 | 110 | 229.82 | 100 | -0.11 | 0.624 |
| Age: | | | | | | | | | | |
| <18 | 361.07 | 75 | 103.01 | 25 | 170.16 | 227.18 | 113.46 | 29 | REF | - |
| 18-60 | 1033.41 | 187.62 | 204.34 | 100 | 328.1 | 110 | 256.15 | 125 | 1.42 | <0.001 |
| >60 | 802.89 | 172 | 230.05 | 133.93 | 272.11 | 69 | 294.5 | 144.93 | 1.18 | 0.011 |
| Occupation: | | | | | | | | | | |
| None | 680.20 | 170.24 | 147.86 | 47.5 | 214.33 | 196.53 | 166.57 | 54.92 | REF | - |
| Casual | 510.00 | 510 | 41.17 | 10 | 0 | 0 | 41.17 | 10 | -1.18 | 0.171 |
| Informal | 930.67 | 161 | 192.86 | 100 | 328.05 | 105 | 246.3 | 120 | -0.09 | 0.781 |
| Formal | 0 | 0 | 236 | 60 | 300 | 300 | 278.86 | 80 | -0.79 | 0.37 |
| Accessed care with NHIS: | | | | | | | | | | |
| No | 419.1 | 95 | 186.84 | 99.09 | 114.89 | 40 | 203.55 | 99.09 | REF | - |
| Yes | 1062.93 | 340 | 169.88 | 70 | 382.71 | 196.53 | 220.2 | 95 | 0.45 | 0.077 |
| Wealth quintile: | | | | | | | | | | |
| Poorest | 841.92 | 177.5 | 179.53 | 70 | 278.05 | 196.53 | 221.43 | 95 | REF | - |
| Poorer | 740.35 | 170.24 | 137.92 | 63.73 | 231.64 | 50 | 182.96 | 80.23 | -0.05 | 0.884 |
| Middle | 1090.67 | 800 | 142.29 | 60 | 577.22 | 326.67 | 189.74 | 60 | -0.60 | 0.096 |
| Richer | 1953.24 | 190.21 | 190.37 | 110 | 182.29 | 125 | 221.18 | 110 | -0.26 | 0.471 |
| Richest | 218.88 | 108 | 225.73 | 96.14 | 419.19 | 231.57 | 260.66 | 99.09 | 0.12 | 0.745 |
| Number of visits: | | | | | | | | | | |
| <4 visits | 598.74 | 100 | 78.83 | 20 | 131.5 | 90 | 85.34 | 20 | REF | - |
| 4 to 10 visits | 546.79 | 340 | 215.06 | 128 | 366.4 | 312 | 232.11 | 130 | 1.19 | <0.001 |
| >10 | 1646.01 | 1600 | 306.49 | 180 | 754.18 | 312 | 413.17 | 230 | 1.94 | <0.001 |
| Constant | - | | - | | | | | | 2.36 | <0.001 |
| R-squared | - | | - | | | | | | 0.2 | - |
| F-test | - | | - | | | | | | 6.66 | <0.001 |
| N | 49 | | 361 | | 49 | | 361 | | 361 | |
| **Total:** | **826.42** | **170.24** | **175.05** | **75** | **295.25** | **112** | **215.12** | **98.19** | **-** | |

* Adjusted coefficients are from a semi-log regression of the association between total financial cost and patient characteristics.

Supplementary Table 5: Total patient costs, by visit time point and cost type

| **Cost type** | **Cost category** | **n** | **Mean** | **Median** | **25th percentile** | **Interquartile** | **75th percentile** | **Minimum** | **Maximum** | **Zero cost** | **Missing** | Toral N (n+Missing) |
| --- | --- | --- | --- | --- | --- | --- | --- | --- | --- | --- | --- | --- |
| ***Before index visit*** | | | | | | | | | | | | |
| Medical expenditure | **Sub-total** | **43** | **581.35** | **100** | **6** | **494** | **500** | **0** | **10000** | **9** | **6** | **49** |
|  | Consultation | 41 | 339.51 | 0 | 0 | 50 | 50 | 0 | 10000 | 27 | 8 | 49 |
|  | Drugs | 39 | 222 | 40 | 0 | 150 | 150 | 0 | 2000 | 17 | 10 | 49 |
|  | Wound dressing | 39 | 62.05 | 0 | 0 | 10 | 10 | 0 | 1000 | 29 | 10 | 49 |
| Non-medical expenditure | **Sub-total** | **41** | **222.15** | **28** | **0** | **200** | **200** | **0** | **3100** | **12** | **8** | **49** |
|  | Food/Accommodation | 40 | 48.77 | 0 | 0 | 5.5 | 5.5 | 0 | 1300 | 29 | 9 | 49 |
|  | Transportation | 40 | 178.93 | 20 | 0 | 137 | 137 | 0 | 1800 | 11 | 9 | 49 |
| **Total financial cost (medical and non-medical) [A]** | **Sub-total** | **43** | **793.16** | **108** | **30** | **770** | **800** | **0** | **10324** | **5** | **6** | **49** |
| **Total financial cost (imputed)** |  | 49 | 826.42 | 170.24 | 50 | 850 | 900 | 0 | 10324 | **4** | **0** | **49** |
| **Productivity loss [B]** | **Sub-total** | **x** | **x** | **x** | **x** | **x** | **x** | **x** | **x** | **x** | **x** | **x** |
| **Total cost [A+B]** | **Total** | **43** | **793.16** | **108** | **30** | **770** | **800** | **0** | **10324** | **5** | **6** | **49** |
| ***During and after index visit (study facility)*** | | | | | | | | | | | | |
| Medical expenditure | **Sub-total** | **334** | **102.84** | **20** | **0** | **120** | **120** | **0** | **1930** | **146** | **27** | **361** |
|  | Consultation | 320 | 26.13 | 0 | 0 | 0 | 0 | 0 | 1930 | 261 | 41 | 361 |
|  | Drugs | 318 | 46.23 | 0 | 0 | 30 | 30 | 0 | 1700 | 206 | 43 | 361 |
|  | Wound dressing | 327 | 34.51 | 0 | 0 | 30 | 30 | 0 | 750 | 227 | 34 | 361 |
| Non-medical expenditure | **Sub-total** | **357** | **72.58** | **20** | **0** | **84** | **84** | **0** | **1440** | **145** | **4** | **361** |
|  | Food/Accommodation | 342 | 5.06 | 0 | 0 | 0 | 0 | 0 | 360 | 304 | 19 | 361 |
|  | Transportation | 355 | 68.12 | 14 | 0 | 80 | 80 | 0 | 1440 | 151 | 6 | 361 |
| **Total expenditure (medical and non-medical) [A]** | **Sub-total** | **358** | **168.32** | **60** | **2** | **208** | **210** | **0** | **3190** | **87** | **3** | **361** |
| **Total cost (imputed)** |  | 361 | 175.05 | 75 | 10 | 200 | 210 | 0 | 3190 | **78** | **0** | **361** |
| **Productivity loss [B]** | **Sub-total** | **361** | **372.72** | **18.16** | **18.16** | **299.64** | **317.8** | **9.08** | **13075.2** | **0** | **0** | **361** |
| **Total cost [A+B]** | **Total** | **361** | **539.64** | **197.12** | **38.16** | **494.24** | **532.4** | **13.62** | **13125.2** | **0** | **0** | **361** |
| ***During and after index visit (non-study facility)*** | | | | | | | | | | | | |
| Medical expenditure | Sub-total | **41** | **154.29** | **50** | **15** | **185** | **200** | **0** | **1080** | **7** | **8** | 49 |
|  | Consultation | 39 | 28.33 | 0 | 0 | 0 | 0 | 0 | 500 | 32 | 10 | 49 |
|  | Drugs | 40 | 80.97 | 10 | 0 | 55 | 55 | 0 | 800 | 19 | 9 | 49 |
|  | Wound dressing | 39 | 50.82 | 0 | 0 | 15 | 15 | 0 | 900 | 27 | 10 | 49 |
| Non-medical expenditure | Sub-total | **42** | **164.48** | **12** | **0** | **100** | **100** | **0** | **1720** | **18** | **7** | 49 |
|  | Food/Accommodation | 41 | 22.93 | 0 | 0 | 0 | 0 | 0 | 750 | 37 | 8 | 49 |
|  | Transportation | 42 | 142.1 | 12 | 0 | 100 | 100 | 0 | 1720 | 18 | 7 | 49 |
| **Total expenditure (medical and non-medical) [A]** | **Sub-total** | **42** | **315.1** | **105** | **30** | **282** | **312** | **0** | **2140** | **5** | **7** | **49** |
| **Total cost (imputed)** |  | 49 | 295.25 | 112 | 30 | 282 | 312 | 0 | 2140 | **5** | **0** | **49** |
| **Productivity loss [B]** | **Sub-total** | x | x | x | x | x | x | x | x | **x** | **x** | **x** |
| **Total cost [A+B]** | **Total** | **42** | **315.1** | **105** | **30** | **282** | **312** | **0** | **2140** | **5** | **7** | **49** |
| ***Combined facilities (study facility and non-study facility)*** | | | | | | | | | | | | |
| Medical expenditure | **Sub-total** | **336** | **121.05** | **27.5** | **0** | **135** | **135** | **0** | **1930** | **139** | **25** | **361** |
|  | Consultation | 324 | 29.22 | 0 | 0 | 0 | 0 | 0 | 1930 | 249 | 35 | 359 |
|  | Drugs | 324 | 55.37 | 0 | 0 | 42.5 | 42.5 | 0 | 1700 | 186 | 35 | 359 |
|  | Wound dressing | 329 | 40.33 | 0 | 0 | 40 | 40 | 0 | 930 | 217 | 30 | 359 |
| Non-medical expenditure | **Sub-total** | **357** | **91.93** | **20** | **0** | **100** | **100** | **0** | **1780** | **139** | **4** | **361** |
|  | Food/Accommodation | 345 | 7.74 | 0 | 0 | 0 | 0 | 0 | 750 | 296 | 15 | 360 |
|  | Transportation | 356 | 84.69 | 20 | 0 | 92.5 | 92.5 | 0 | 1780 | 136 | 4 | 360 |
| **Total expenditure (medical and non-medical) [A]** | **Sub-total** | **358** | **205.28** | **70** | **4** | **246** | **250** | **0** | **3190** | **83** | **3** | **361** |
| **Total cost (imputed)** |  | 361 | 215.12 | 98.19 | 10 | 270 | 280 | 0 | 3190 | 73 | 0 | 361 |
| **Productivity loss [B]** | **Sub-total** | **361** | **372.72** | **18.16** | **18.16** | **299.64** | **317.8** | **9.08** | **13075.2** | **0** | **0** | **361** |
| **Total cost [A+B]** | **Total** | **361** | **576.3** | **198.16** | **38.16** | **555.04** | **593.2** | **13.62** | **13187.2** | **0** | **0** | **361** |

Supplementary Figure 1: Map of study site (Atwima Mponua district) within Republic of Ghana (upper panel) and at district level (lower panel)

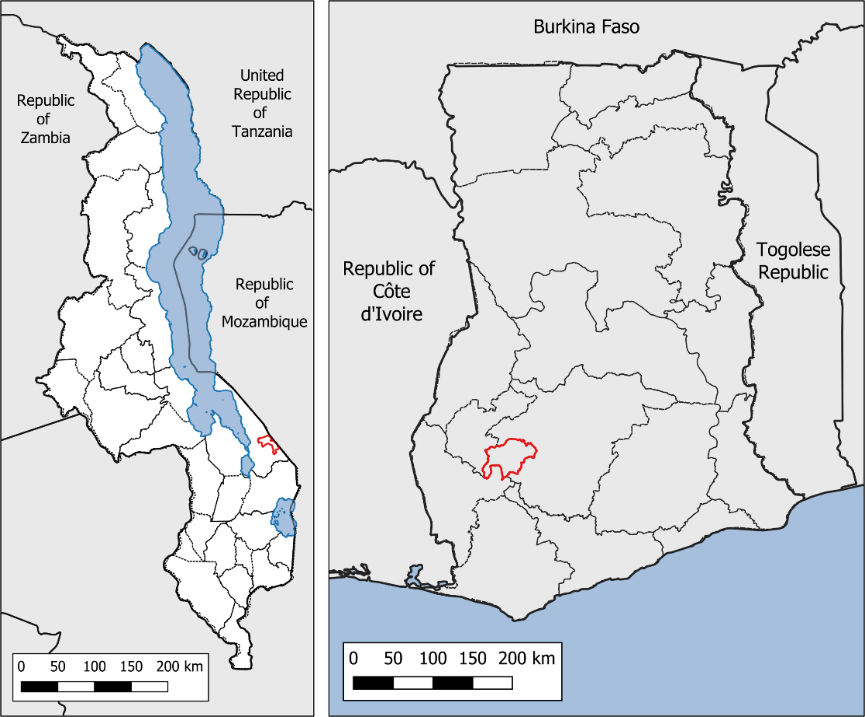

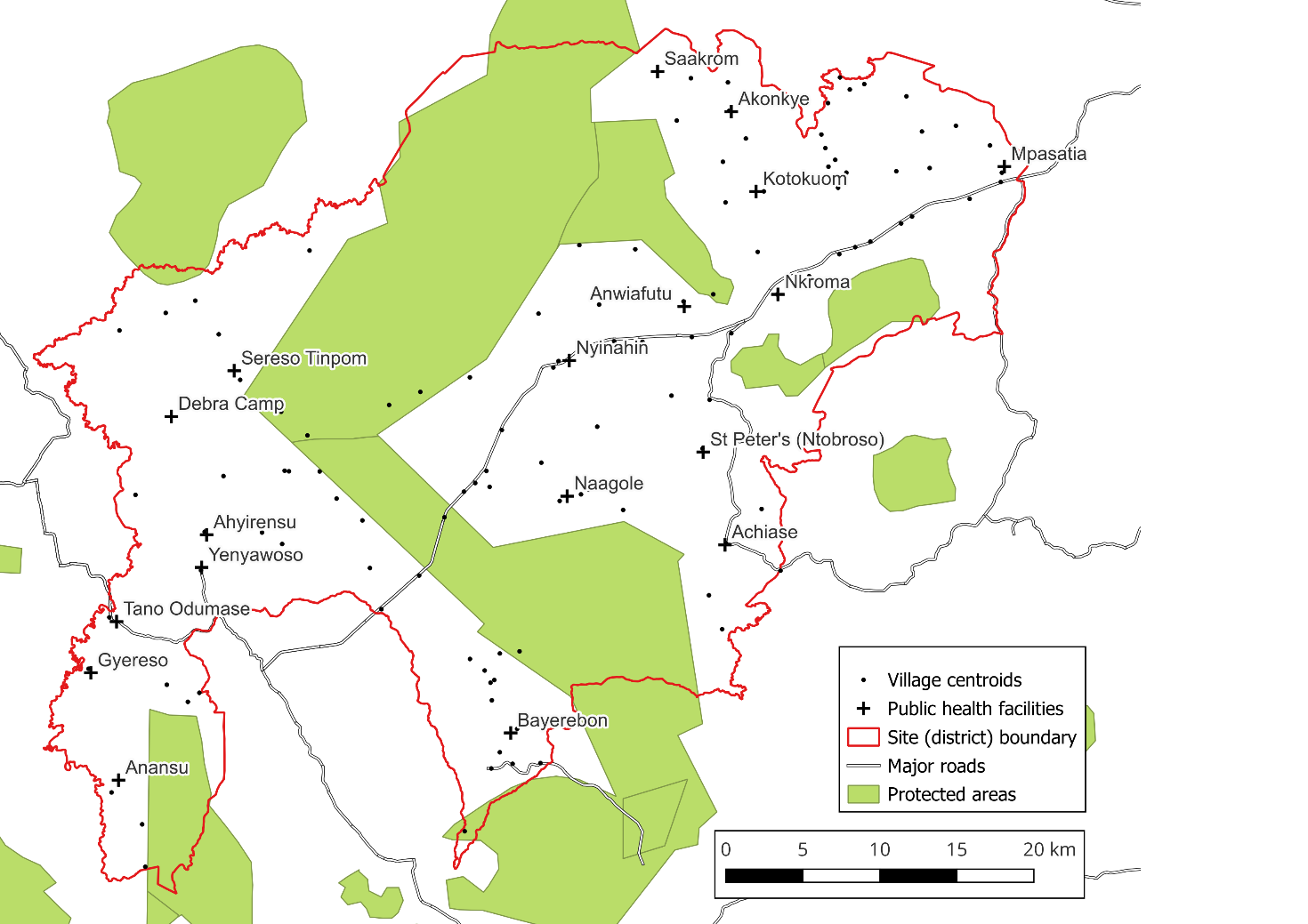

Supplementary Figure 2: Association between community-level incidence and endline prevalence

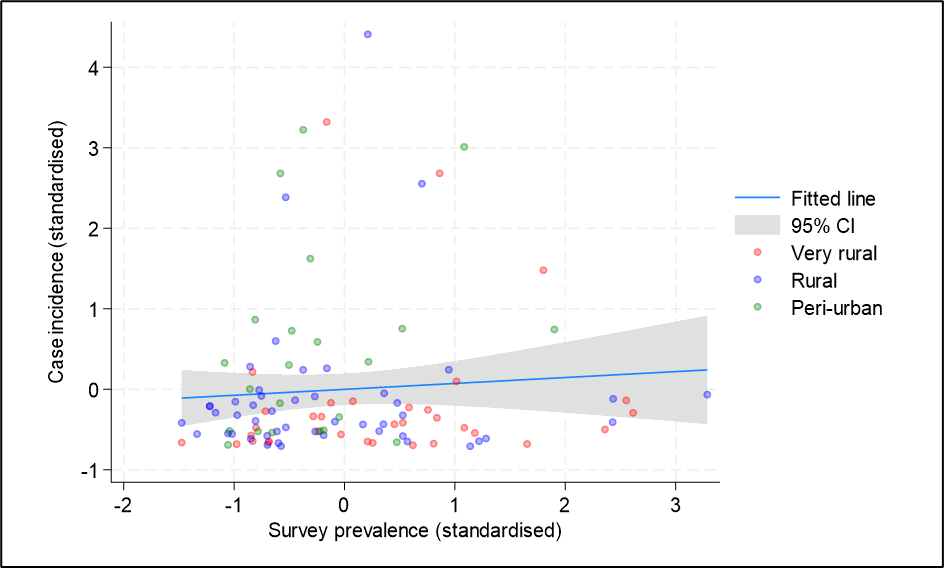

Scatter plot of standardised community-level prevalance of skin conditions at endline survey (July–September 2024) versus standardised community-level incidence of skin conditions (November 2023–September 2024) amongst endline survey communities. R^2^ (coefficient of determination)=0.006

Supplementary Figure 3: Provider costs, by health facility and resource category

Ghana Cedis (2024)

|  | Consumables | Human resources | Combined transport (vehicle-fuel-reimbursement) | Capital |
| --- | --- | --- | --- | --- |
| CHPS HF5 | 28831.9 | 14098 | 5639.64 | 1824.94 |
| CHPS HF1 | 29547.9 | 15141 | 5813.94 | 2373.23 |
| CHPS HF11 | 30690.4 | 18233 | 5636.03 | 2228.38 |
| CHPS HF8 | 38810.5 | 20119 | 8704.51 | 4044.95 |
| CHPS HF6 | 42723.1 | 19900.4 | 9075.23 | 2465.04 |
| CHPS HF10 | 41613.2 | 27886.1 | 12071.6 | 4455.45 |
| CHPS HF3 | 46376.2 | 29677.9 | 8584.94 | 2924.37 |
| Health Center HF13 | 17614 | 8893.52 | 2773.57 | 4393.71 |
| Health Center HF15 | 37883.5 | 20372.3 | 6769.41 | 3481.2 |
| Health Center HF9 | 37019.6 | 19606.4 | 8477.39 | 3536.22 |
| Health Center HF16 | 38643.7 | 21830.5 | 6720.56 | 3962.85 |
| Health Center HF12 | 45377.6 | 23384.7 | 9933.4 | 3837.22 |
| Health Center HF7 | 48846.6 | 23229 | 8691.11 | 2610.35 |
| Health Center HF14 | 75794.9 | 36751.1 | 13316.7 | 4532.67 |
| Health Center HF2 | 77413.8 | 45138.1 | 14222.5 | 4525.23 |
| Health Center HF4 | 109796 | 49611.9 | 20157.2 | 6168.1 |
| DH HF17 | 106579 | 47966.8 | 29757.2 | 10569.3 |

Supplementary Figure 4: Provider costs, by activity and resource category

Ghana Cedis (2024)

|  | Consumables | Human resources | Combined transport (vehicle-fuel-reimbursement) | Capital |
| --- | --- | --- | --- | --- |
| Clinical care | 540752 | 49271.6 | 17961.4 | 36828.8 |
| Training | 112950 | 214320 | 140430 | 8200 |
| Community engagement | 192559 | 59328 | 7453.49 | 9954.38 |
| Preparation | 4800 | 109020 |  | 12450 |
| M&E | 2500 | 9900 | 10500 | 500 |
